# Short and long-term prognosis after tissue negative transient ischemic attack

**DOI:** 10.1101/2024.02.11.24302667

**Authors:** Francisco Purroy, Yhovany Gallego, M Pilar Gil-Villar, Robert Begue, Gloria Arque, Alejandro Quilez, Jordi Sanahuja, Daniel Vazquez-Justes, Gerard Mauri

## Abstract

**Background:** Recently, there has been a proposal to retire the concept of transient ischemic attack (TIA) because, in all likelihood, patients experiencing brief episodes of transient brain ischemia without associated cerebral lesions might be exceedingly rare or carry no risk of stroke recurrence (SR). However, only a few observational studies have evaluated the risk of SR exclusively among tissue-negative TIA patients. Our aim was to assess the early and long-term prognosis of consecutive tissue-negative TIA patients attended at an emergency department

**Methods:** We carried out a prospective cohort study of consecutive TIA patients with tissue negative TIA from January 2006 to June 2010. All patients underwent diffusion-weighted imaging on MRI (DWI) (4.0 [SD 1.8] days) after the index event. The risk and predictors of SR were determined at 1 year and after a median follow-up time of 6.6 (interquartile range, 5.0-9.6) years.

**Results:** A total of 370 patients were included. Previously, 244 patients with positive DWI results and 109 patients without MRI performed were excluded. ABCD2 score>5 was determined in 95 (26.2%) patients. 15 (4.1%) patients suffered SR at 1 year and 18 (4.9%) beyond 1 year. Predictive models for short-term and long-term prognosis were different. Large artery atherosclerosis etiology (Hazard ratio [HR] 3.7 [1.2-11.0]) was the only predictor of 1 year SR. In contrast, male sex (HR 4.17 [95% CI 1.14-15.23]; P=0.031), speech impairment (HR 4.90 [95% CI 1.05-22.93]; P=0.044), and presence of chronic microangiopathy expressed as Fazekas score of 3 (HR 1.84 [95% CI 1.15-2.97]; P=0.012) were predictors of long-term SR follow-up.

**Conclusion:** The risk of SR after tissue negative TIA is not insignificant. Predictors of short and long-term prognosis are different. Sex, clinical characteristics at onset, etiology and chronic microangiopathy determine the risk of SR.

## INTRODUCTION

Since the tissue definition of transient ischemic attack (TIA) was established, TIA has been defined as a transient episode of neurological symptoms without evidence of brain infarction^1^. Recently, Recently, there has been a proposal to retire this concept because it is likely that, with the improvement of neuroimaging techniques, patients experiencing brief episodes of transient brain ischemia actually have lesions in the cerebral parenchyma. It is suggested that these patients should be considered stroke patients. Additionally, it is argued that individuals who suffer transient ischemia without any lesions either do not exist or, if they do, are rare and pose no risk of stroke recurrence (SR) ^2^. Until the implementation of imaging tests more sensitive to acute ischemia, it is compelling to investigate the SR in diffusion-weighted imaging (DWI) negative patients. However, no observational studies have exclusively determined the risk of stroke recurrence among tissue-negative TIA patients. Large studies that included both tissue-positive and tissue-negative patients identified predictors of early and late stroke recurrence beyond neuroimaging data, such as large-artery atherosclerosis, clinical features, and sex differences^3-5^.

Our aim was to assess the early and long-term prognosis of consecutive tissue-negative TIA patients who were attended at an emergency department and to identify predictors of SR among these patients.

## METHODS

### Data availability statement

The corresponding author will consider requests for access to the data reported in this article.

### Design and study population

The methodology of the study has been previously described^4^. Consecutive patients were included from the cohort of patients in the REGITELL registry)^6^. REGITELL registry is a registry-based cohort study conducted from January 2006 to June 2010, involving consecutive TIA patients attended by a stroke neurologist working in the emergency department of a university hospital within the first 24 hours after the onset of symptoms. The registry was designed in compliance Strengthening the Reporting of Observational Studies in Epidemiology (STROBE) statement.^7^. We included patients who experienced a reversible episode of neurological deficit of ischemic origin that completely resolved within 24 hours and exhibited no evidence of acute ischemic lesions in diffusion-weighted imaging (DWI) sequences ^1^. In all cases, cranial MRI was conducted within 7 days of symptom onset (4.0[standard deviation, SD 1.8] days)^4,6^. Patients were excluded if they had contraindications for cranial MRI or if the procedure could not be performed. The clinical symptom typologies were documented using a standardized form. The symptom duration was confirmed upon admission and reverified on day 7 during the follow-up period. Their respective ABCD2 risk scores (age, blood pressure, clinical features, symptom duration, and diabetes mellitus)^8^ were prospectively assessed. Possible but not conclusively diagnosed TIA events were characterized by isolated atypical symptoms, including unsteadiness, diplopia, dysarthria, partial sensory deficit, and unusual cortical vision^9^. Every case was reviewed and prospectively classified by the senior neurologist (FP). Patients underwent initial investigations, including ECG, blood tests, CT brain imaging, and extracranial and intracranial ultrasound imaging^6^. Patients in whom cardiopathy was suspected underwent comprehensive cardiological examinations. The etiological classification of patients was conducted according to the definitions outlined in the ORG 10172 Trial (TOAST).^10^ *Imaging Analysis*

Alongside DWI sequences, we incorporated T2-weighted turbo spin-echo, T2-weighted gradient-recalled echo (T2*-GRE), T1-weighted spin-echo, and turbo fluid-attenuated inversion recovery sequences ^6^. A neuroradiologist blinded (RB) to the patients’ clinical data determined the presence of white matter lesions using FLAIR sequences on the MRI. He assessed the Fazekas scale score by evaluating both periventricular and deep WM alterations, considering the quantity and size of lesions, and distinguishing between focal or punctate, early confluent, and confluent abnormalities, scoring from 0 (absence of WM lesions) to 3 (presence of diffuse WM lesions)^11^. Additionally, it ascertained the presence of chronic ischemic lesions in FLAIR sequences corresponding to territorial infarcts not suggestive of microangiopathy. The presence of cerebral microbleeds (CMBs) was determined using T2*-GRE. According to consensus criteria, CMBs were defined as homogeneous hypointense lesions located in the gray or white matter, distinguishable from iron or calcium deposits and vessel flow voids ^12^.

### Outcomes and follow-up

The primary outcome was the incidence of recurrent ischemic stroke, characterized by the emergence of new focal symptoms or signs accompanied by acute ischemic changes detected on neuroimaging via brain CT or MRI. Recurrent TIAs, defined as new neurological symptoms or deficits lasting less than 24 hours without evidence of new infarction in neuroimaging, were not considered part of the primary outcome ^4^. The secondary outcome comprised a composite of major vascular events (MVE). This encompassed acute coronary syndrome defined as myocardial infarction (with or without ST-segment elevation) or unstable angina leading to urgent catheterization, recurrent ischemic stroke, the onset of symptomatic peripheral arterial disease (PAD), and cardiovascular-related fatalities.

Cardiovascular deaths included fatal acute coronary syndrome, fatal stroke, fatal intracranial hemorrhage, fatal pulmonary embolism, sudden death, and unexpected or unobserved deaths within 30 days^5^. During the follow-up period, structured clinical visits were conducted by a team of stroke physicians. These visits occurred at specific intervals of 7 days, 3 months, one year, five years, and 10 years. In instances where a patient relocated out of the local area or hospital visits were not feasible, follow-up assessments were carried out via telephone. Moreover, recurrent events and outcomes were actively monitored through an annual review of electronic medical records.

### Statistical analysis

We compared the baseline characteristics, etiology, neuroimaging data, and ABCD2 score^8^ between patients with and without SR. Quantitative variables were compared using either Student’s t-test or the Mann-Whitney U test. Qualitative variables were compared using the chi-squared test or Fisher’s exact test in cases where the expected cell frequency was less than 5. We employed a Cox proportional hazards multivariable analysis, incorporating variables with p-values <0.10 from univariate testing, to identify predictors of SR. The cumulative risks of recurrent stroke and MVE during the follow-up were assessed through Kaplan-Meier analysis. Results were censored at the occurrence of the outcome event, patient death, or the conclusion of the follow-up period. For patients with missing information at the 10-year mark, data were censored at the time of their last available follow-up. We conducted a comparison of the risk of SR among patient subgroups, categorized based on the primary identified predictors, utilizing Kaplan-Meier analysis and the log-rank test. All analyses were two-sided. In multivariable analyses, missing data was accounted for as a random effect when fitting the model. The statistical analysis of the data was carried out using the SPSS statistical package, version 25.0. (IBM SPSS, Chicago, IL, USA) and Graphpad software version 6 (San Diego, CA, USA).

### Standard protocol approvals, registrations, and patient consents

Written informed consent or assent from relatives was obtained for all the participants. The study was approved by our local ethics committee: the “Comité d’Etica i Investigació Clínica de l’Hospital Universitari Arnau de Vilanova de Lleida”.

## RESULTS

A total of 771 patients were initially assessed as possible TIA cases. Forty-eight patients were excluded after being diagnosed as mimics. One hundred nine patients (15.1%) out of the remaining 723 were excluded from the analysis as they did not undergo an MRI. Of the 614 patients who underwent DWI, 370 (60.3%) did not exhibit acute ischemic lesions and were thus included in the study. The median (interquartile rate, IQR) follow-up time was 6.6 (5.2-9.7) years. Thirty-three individuals (8.9%) experienced stroke recurrence: 15 (45.5%) of them during the first year of follow-up and 18 (54.5%) beyond the initial year of follow-up (figure 1). MVE were observed in 47 (12.7%) patients: 22 (46.8%) of them during the first year of follow-up and 25 (53.2%) beyond the initial year of follow-up (figure 1). A total of 111 (30.0) patients died during the follow-up. The median Fazekas score was 0 (IQR: 0-2.0), and an ABCD2 score >5 was present in 95 patients (25.7%)

**FIGURE 1.**
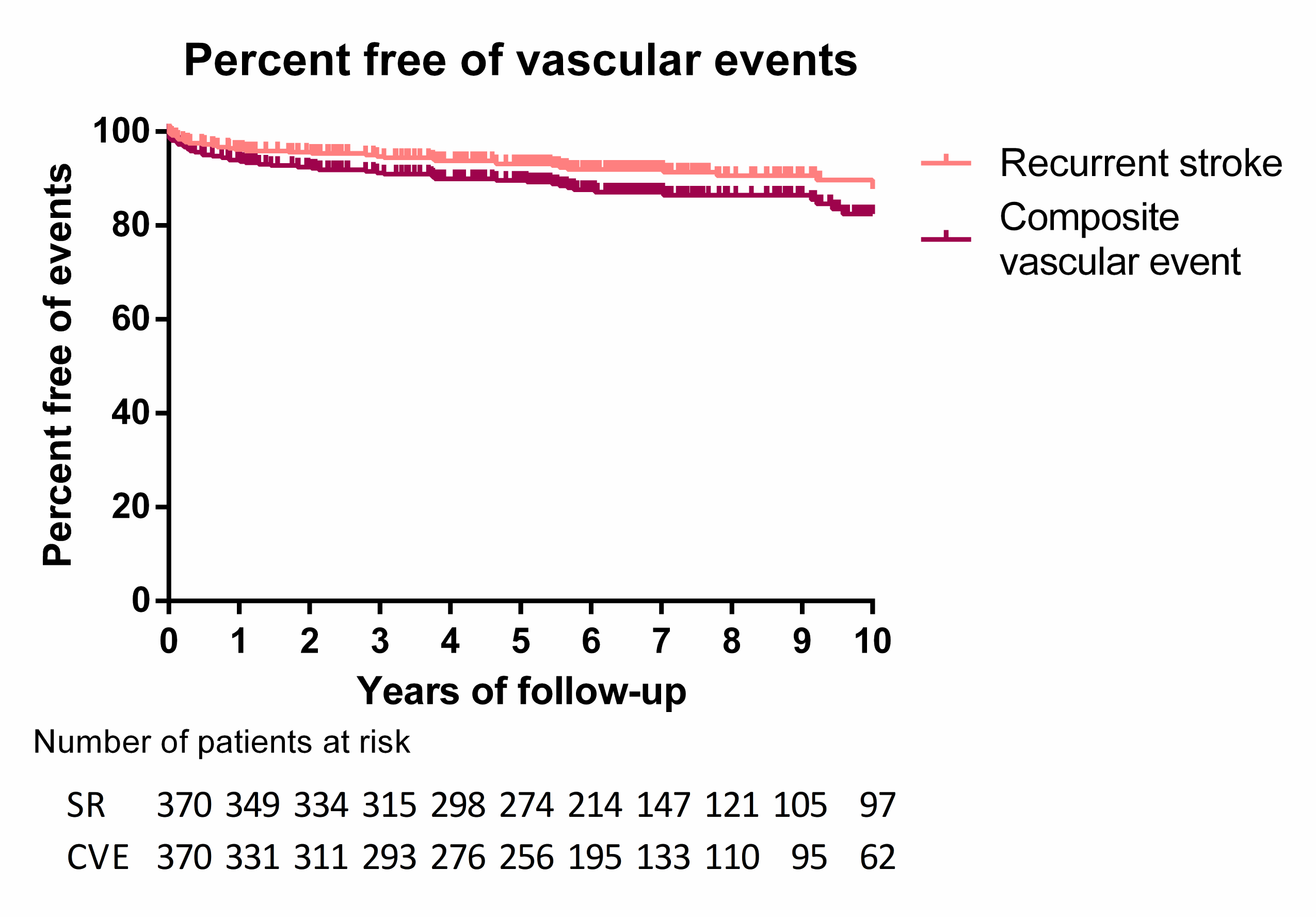
Kaplan-Meier event curves for stroke recurrence and major vascular events according along the entire follow-up.

### Variables associated with stroke recurrence

As depicted in Table 1, various variables are differently associated with early SR within the first year of follow-up and with the risk of SR from the subsequent year of follow-up. Older patients (75.5 [SD: 7.1] vs. 70.2 [SD: 12.1] years; P=0.093), patients with previous atrial fibrillation (11.1% vs. 3.3%; P=0.024) and those with a large artery atherosclerosis subtype etiology (10.2% vs. 2.9%; P=0.009). were associated with early SR within the first year of follow-up. Conversely, patients with diabetes mellitus (9.8% vs. 3.3%; P=0.021), ABCD2 score >5 (12.3% vs. 3.1%; P=0.003), and a Fazekas score of 3 (11.0% vs. 3.7%; P=0.057) were associated with SR from the subsequent year of follow-up. In the subsequent year of follow-up, women had a lower risk of stroke (2.7% vs. 7.2%; P=0.076). Ultimately, patients experiencing speech impairment during the index event were associated with stroke recurrence throughout the entire follow-up period (5.6% vs. 1.4%; P=0.050 and 7.5% vs. 1.5%; P=0.020).

**Table 1.**
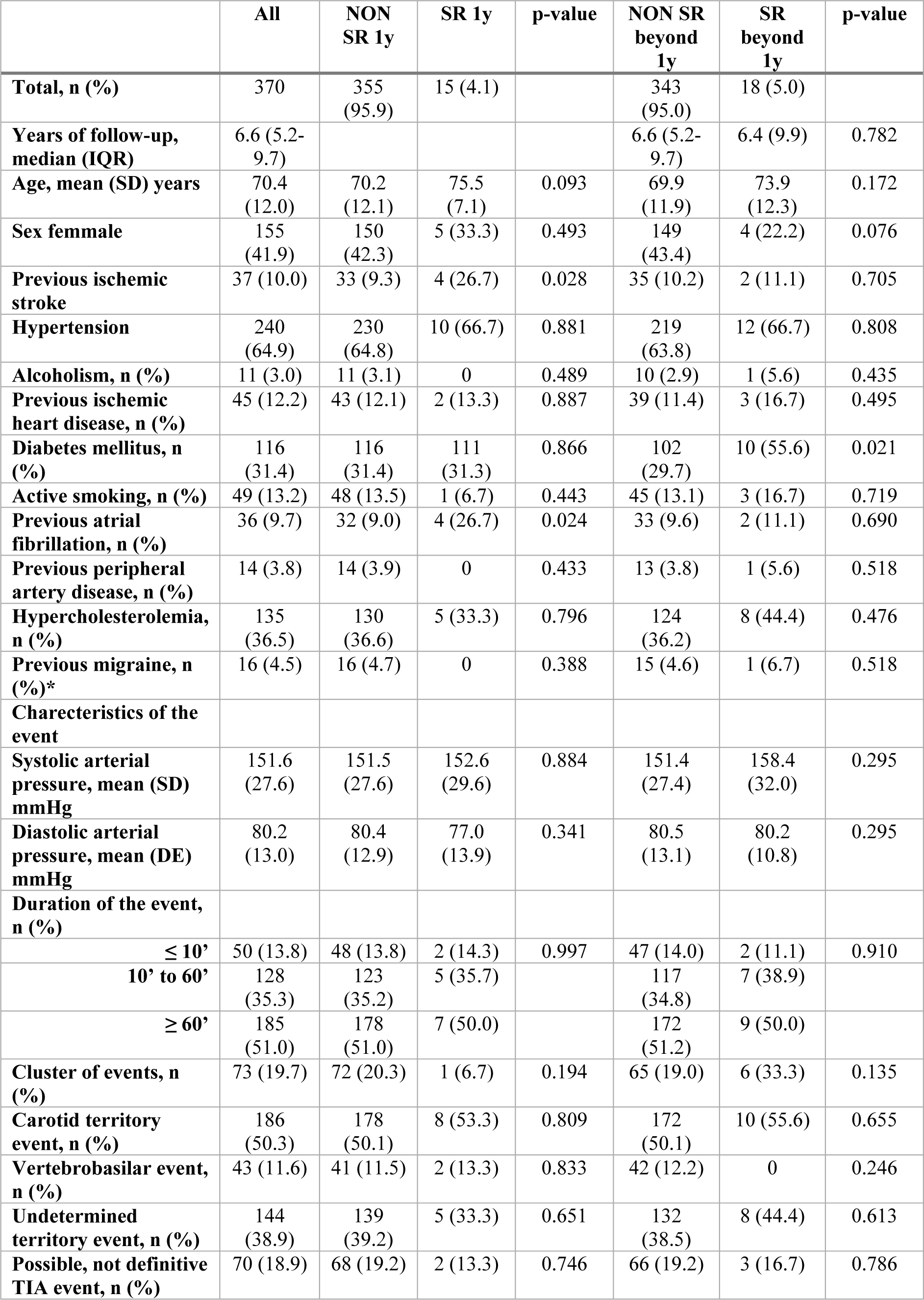
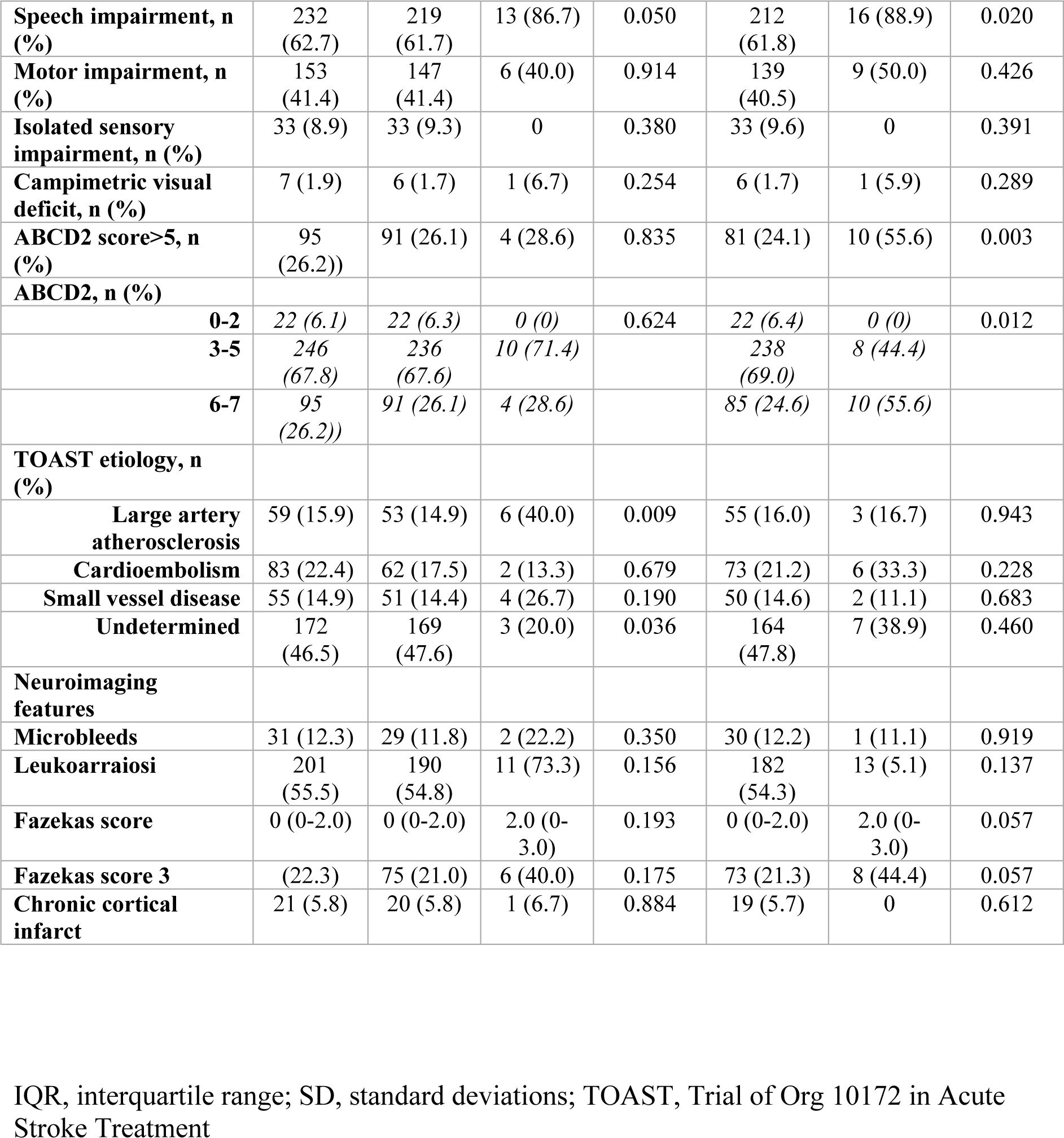
Baseline characteristics, etiology and neuroimaging findings by stroke recurrence.

### Predictors of stroke recurrence

SR during the first year of follow-up was significantly associated with large artery atherosclerosis etiology (hazard ratio [HR] 3.67 [95% CI, 1.22–11.04]; P=0.021). Conversely, stroke recurrence beyond the initial year of follow-up was linked to male sex (HR 4.17 [95% CI 1.14-15.23]; P=0.031), speech impairment (HR 4.90 [95% CI 1.05-22.93]; P=0.044), and a Fazekas score of 3 (HR 3.36 [95% CI 1.12-10.11]; P=0.031) (Table 2).

**Table 2.** Follow-up events.

| Follow-up events | All | One year of follow-up | Beyond one year of follow-up |
| --- | --- | --- | --- |
| TIA recurrence, n (%) | 50 (13.5) | 33 (8.9) | 17 (4.7) |
| Acute coronary syndrome, n (%) | 12 (3.2) | 6 (1.6) | 6 (1.7) |
| Peripheral artery disease, n (%) | 6 (1.6) | 3 (0.8) | 3 (0.8) |
| Death, n (%) | 111 (30.0) | 13 (3.5) | 98 (27.1) |
| Vascular death, n (%) | 9 (2.4) | 6 (1.6) | 3 (0.8) |
| Composite vascular endpoint, n (%) | 47 (12.7) | 22 (5.9) | 25 (6.9) |

**Table 3.** Cox proportional hazards regression model to assess risk of subsequent stroke after transient ischemic attack, considering time-periods of follow-up.

|  | <= 1 year |  | > 1 year |  |
| --- | --- | --- | --- | --- |
|  | HR (95% CI) | p-value | HR (95% CI) | p-value |
| Age | 1.02 (0.96-1.08) | 0.504 | - | - |
| Sex male | - | - | 4.17 (1.14-15.23) | 0.031 |
| Diabetes mellitus | - | - | 0.95 (0.27-3.34) | 0.942 |
| Previous ischemic stroke | 2.16 (0.64-7.25) | 0.212 | - | - |
| Speech impairment | 3.38 (0.75-15.24) | 0.113 | 4.90 (1.05-22.93) | 0.044 |
| Previous atrial fibrillation | 3.51 (1.00-12.41) | 0.212 | - | - |
| ABCD2>5 | - | - | 2.00 (0.60-6.70) | 0.261 |
| Large artery atherosclerosis etiology | 3.67 (1.22-11.04) | 0.021 | - | - |
| Fazekas score of 3 | - | - | 3.36 (1.12-10.11) | 0.031 |

In the Kaplan-Meier analysis (Figure 2), an undetermined etiology was associated with the lowest risk of stroke recurrence throughout the follow-up period, whereas LAA was associated with a higher risk of stroke recurrence during the first year (P=0.077). The risk of SR trend to remained elevated in those patients with a Fazekas score of 3 from the first to the second year of follow-up (P=0.051). Similarly, patients with ABCD2 scores >5 maintained a heightened risk, especially from the second year of follow-up onwards (P=0.013). The risk of SR remained similar between both sexes during the first three years, later tending to be higher in male patients (P=0.051). Finally, patients with speech impairment have a higher risk of stroke recurrence throughout the entire follow-up (P=0.001).

**FIGURE 2.**
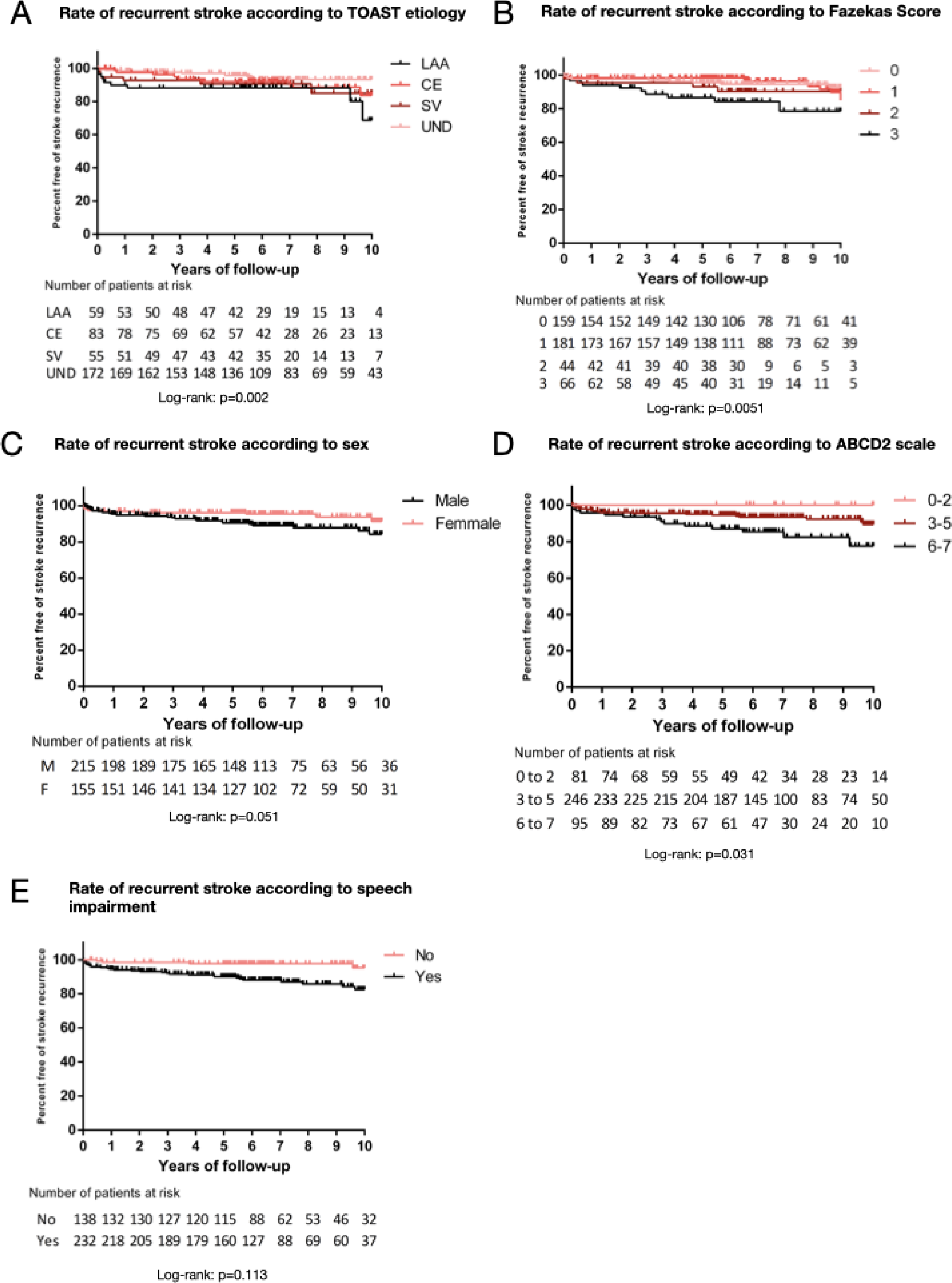
Kaplan-Meier event curves for stroke recurrence according to TOAST etiology (A), Fazekas scale (B), sex (C), ABCD2 score (D) and speech impairment (E)

## DISCUSSION

In a cohort of patients with TIA, based on tissue definition, we observed that the risk of stroke recurrence, although lower than expected for patients with minor stroke or abnormal DWI TIA^3-5,13^, is not negligible, affecting nearly one in ten patients. Thus, the underestimation of the risk of SR in those patients with episodes compatible with TIA and normal DWI, as well as the trivialization of the concept of TIA, needs to be approached with caution^2^. According to our results, it is essential to consider that there are variables that determine the risk of recurrence differently throughout the follow-up. In our study, similar to previous research that included patients with transient clinical symptoms regardless of DWI results, as well as those with minor strokes, atheromatous etiology^3-5^ was linked to more than threefold increase in the risk of stroke recurrence during the first year. Beyond the one-year follow-up, non-modifiable risk factors such as gender, the presence of chronic microangiopathy, and certain clinical characteristics identified at the time of the episode become relevant. In line with previous studies, we observed a higher recurrence risk among male patients^4,14-16^. These differences could be attributed to a higher accumulation of vascular risk factors among men compared to women^4,16^. Interestingly, we observed an association between the presence of white matter hyperintensities (WMH) or leukoaraiosis, particularly when they are extensive and reach Fazekas grade 3, and SR. Previously, it has been described how the burden of chronic white matter lesions was associated with short-term outcomes^17^ and long-term SR^18^. WMH are a common neuroimaging feature of cerebral microangiopathy^19^. A higher rate of extended WMH is strongly associated with SR in all stroke subtypes, even after adjustment for clinical risk factors^20^. Some previous studies have also been focused on TIA patients. Ren et al. demonstrated a correlation between WMH and SR, which can be attributed to arteriosclerotic narrowing, plaque, artery stiffening, small vessel disease, hemodynamic injury, and impaired autoregulation^21^. Furthermore, we observed the relevance of the clinical characteristics of the index episode in identifying patients at higher risk of recurrence. Thus, there was an association between high scores on the ABCD2 scale and SR during advanced follow-up. Unlike TIA studies that do not consider tissue definition, language impairment has assumed a more prominent role than motor impairments or the presence of multiple episodes in our investigation^22^. It is possible that these characteristics are linked to a higher likelihood of acute ischemic lesions on DWI^23^. Hence, such patients would be excluded in our series. It can be highlighted that none of the patients with isolated sensory symptoms experienced a SR.

Our study has certain limitations that need to be highlighted. On one hand, it is essential to consider that this is a single-center study, which poses a limitation in terms of sample size. On the other hand, we must acknowledge that the analysis of WMH has been qualitative rather than quantitative, although there are studies that have addressed both approaches simultaneously without significant differences between them^17,24^.

The risk of SR following a tissue-negative TIA is noteworthy. Predictors for short and long-term prognosis vary, with sex, initial clinical characteristics, etiology and WMH being determinants of the SR risk.

The risk of SR after a tissue-negative TIA is of significant concern. Predictors influencing short and long-term prognosis are multifaceted, including sex, initial clinical characteristics, etiology, and the presence and extension of WMH, all of which serve as determining factors for the SR risk at different stages during follow-up. Thus, there should be a reinforcement of the therapeutic and organizational management for patients who have experienced a tissue-negative TIA of atheromatous etiology to prevent recurrences during the initial year of follow-up. Additionally, for those patients exhibiting elevated scores on the ABCD2 scale and extensive white matter lesions, particular emphasis should be placed on averting long-term recurrences.

## Contributions

FP conceived the study and procured the funding

FP, YG, MPGV, GM, AQ, JS, DVJ, were involved in the recruitment and follow-up of patients

FP wrote the paper. FP and GM substantively revised the manuscript

All the authors revised and approved the final manuscript

## Non-standard Abbreviations and Acronyms

AF: atrial fibrillation
DWI: diffusion weighted imaging
HR: hazard ratio
LAA: large artery atherosclerosis
MVE: major vascular events
SR: stroke recurrence
WMH: white matter hyperintensities

## REFERENCES

1. Easton JD, Saver JL, Albers GW, Alberts MJ, Chaturvedi S, Feldmann E, Hatsukami TS, Higashida RT, Johnston SC, Kidwell CS, et al. Definition and evaluation of transient ischemic attack: a scientific statement for healthcare professionals from the American Heart Association/American Stroke Association Stroke Council; Council on Cardiovascular Surgery and Anesthesia; Council on Cardiovascular Radiology and Intervention; Council on Cardiovascular Nursing; and the Interdisciplinary Council on Peripheral Vascular Disease. The American Academy of Neurology affirms the value of this statement as an educational tool for neurologists. Stroke. 2009;40:2276–2293. doi: 10.1161/strokeaha.108.192218

2. Easton JD, Johnston SC. The Concept of Transient Ischemic Attack-Reply. Jama. 2022;327:2457. doi: 10.1001/jama.2022.7630

3. Ois A, Zabalza A, Moreira A, Cuadrado-Godia E, Jimenez-Conde J, Giralt-Steinhauer E, Rodriguez-Campello A, Soriano-Tarraga C, Roquer J. Long-term cardiovascular prognosis after transient ischemic attack: Associated predictors. Neurology. 2018;90:e553–e558. doi: 10.1212/WNL.0000000000004965

4. Purroy F, Vicente-Pascual M, Arque G, Baraldes-Rovira M, Begue R, Gallego Y, Gil MI, Gil-Villar MP, Mauri G, Quilez A, et al. Sex-Related Differences in Clinical Features, Neuroimaging, and Long-Term Prognosis After Transient Ischemic Attack. Stroke. 2021;52:424–433. doi: 10.1161/STROKEAHA.120.032814

5. Amarenco P, Lavallee PC, Monteiro Tavares L, Labreuche J, Albers GW, Abboud H, Anticoli S, Audebert H, Bornstein NM, Caplan LR, et al. Five-Year Risk of Stroke after TIA or Minor Ischemic Stroke. N Engl J Med. 2018;378:2182–2190. doi: 10.1056/NEJMoa1802712

6. Purroy F, Begue R, Gil MI, Quilez A, Sanahuja J, Brieva L, Pinol-Ripoll G. Patterns of diffusion-weighted magnetic resonance imaging associated with etiology improve the accuracy of prognosis after transient ischaemic attack. Eur J Neurol. 2011;18:121–128. doi: ENE3080 [pii] 10.1111/j.1468-1331.2010.03080.x

7. von Elm E, Altman DG, Egger M, Pocock SJ, Gotzsche PC, Vandenbroucke JP, Initiative S. The Strengthening the Reporting of Observational Studies in Epidemiology (STROBE) statement: guidelines for reporting observational studies. Lancet. 2007;370:1453–1457. doi: 10.1016/S0140-6736(07)61602-X

8. Johnston SC, Rothwell PM, Nguyen-Huynh MN, Giles MF, Elkins JS, Bernstein AL, Sidney S. Validation and refinement of scores to predict very early stroke risk after transient ischaemic attack. Lancet. 2007;369:283–292.

9. Lavallee PC, Sissani L, Labreuche J, Meseguer E, Cabrejo L, Guidoux C, Klein IF, Touboul PJ, Amarenco P. Clinical Significance of Isolated Atypical Transient Symptoms in a Cohort With Transient Ischemic Attack. Stroke. 2017;48:1495–1500. doi: 10.1161/STROKEAHA.117.016743

10. Adams HP, Jr., Bendixen BH, Kappelle LJ, Biller J, Love BB, Gordon DL, Marsh EE, 3rd. Classification of subtype of acute ischemic stroke. Definitions for use in a multicenter clinical trial. TOAST. Trial of Org 10172 in Acute Stroke Treatment. Stroke. 1993;24:35-41.

11. Wardlaw JM, Smith EE, Biessels GJ, Cordonnier C, Fazekas F, Frayne R, Lindley RI, O’Brien JT, Barkhof F, Benavente OR, et al. Neuroimaging standards for research into small vessel disease and its contribution to ageing and neurodegeneration. Lancet Neurol. 2013;12:822–838. doi: 10.1016/S1474-4422(13)70124-8

12. Greenberg SM, Vernooij MW, Cordonnier C, Viswanathan A, Al-Shahi Salman R, Warach S, Launer LJ, Van Buchem MA, Breteler MM, Microbleed Study G. Cerebral microbleeds: a guide to detection and interpretation. Lancet Neurol. 2009;8:165–174. doi: 10.1016/S1474-4422(09)70013-4

13. Shahjouei S, Sadighi A, Chaudhary D, Li J, Abedi V, Holland N, Phipps M, Zand R. A 5-Decade Analysis of Incidence Trends of Ischemic Stroke After Transient Ischemic Attack: A Systematic Review and Meta-analysis. JAMA Neurol. 2021;78:77–87. doi: 10.1001/jamaneurol.2020.3627

14. Yu AYX, Penn AM, Lesperance ML, Croteau NS, Balshaw RF, Votova K, Bibok MB, Penn M, Saly V, Hegedus J, et al. Sex Differences in Presentation and Outcome After an Acute Transient or Minor Neurologic Event. JAMA neurology. 2019;76:962–968. doi: 10.1001/jamaneurol.2019.1305

15. Madsen TE, Howard VJ, Jimenez M, Rexrode KM, Acelajado MC, Kleindorfer D, Chaturvedi S. Impact of Conventional Stroke Risk Factors on Stroke in Women: An Update. Stroke. 2018. doi: 10.1161/STROKEAHA.117.018418

16. Cordonnier C, Sprigg N, Sandset EC, Pavlovic A, Sunnerhagen KS, Caso V, Christensen H, Women Initiative for Stroke in Europe g. Stroke in women - from evidence to inequalities. Nat Rev Neurol. 2017;13:521–532. doi: 10.1038/nrneurol.2017.95

17. Zerna C, Yu AYX, Modi J, Patel SK, Coulter JI, Smith EE, Coutts SB. Association of White Matter Hyperintensities With Short-Term Outcomes in Patients With Minor Cerebrovascular Events. Stroke. 2018;49:919–923. doi: 10.1161/STROKEAHA.117.017429

18. Kumral E, Gulluoglu H, Alakbarova N, Karaman B, Deveci EE, Bayramov A, Evyapan D, Gokcay F, Orman M. Association of leukoaraiosis with stroke recurrence within 5 years after initial stroke. J Stroke Cerebrovasc Dis. 2015;24:573–582. doi: 10.1016/j.jstrokecerebrovasdis.2014.10.002

19. Wardlaw JM, Smith C, Dichgans M. Small vessel disease: mechanisms and clinical implications. Lancet Neurol. 2019;18:684–696. doi: 10.1016/S1474-4422(19)30079-1

20. Dimaras T, Merkouris E, Tsiptsios D, Christidi F, Sousanidou A, Orgianelis I, Polatidou E, Kamenidis I, Karatzetzou S, Gkantzios A, et al. Leukoaraiosis as a Promising Biomarker of Stroke Recurrence among Stroke Survivors: A Systematic Review. Neurol Int. 2023;15:994–1013. doi: 10.3390/neurolint15030064

21. Ren XM, Qiu SW, Liu RY, Wu WB, Xu Y, Zhou H. White Matter Lesions Predict Recurrent Vascular Events in Patients with Transient Ischemic Attacks. Chin Med J (Engl). 2018;131:130–136. doi: 10.4103/0366-6999.222341

22. Purroy F, Jiménez-Caballero PE, Gorospe A, Torres MJ, Álvarez-Sabín J, Santamarina E, Martínez-Sánchez P, Cánovas D, Freijó MM, Egido JA, et al. Recurrent transient ischaemic attack and early risk of stroke: data from the PROMAPA study. J Neurol Neurosurg Psychiatry. 2013;84:596–603.

23. Purroy F, Begue R, Quilez A, Pinol-Ripoll G, Sanahuja J, Brieva L, Seto E, Gil MI. The California, ABCD, and unified ABCD2 risk scores and the presence of acute ischemic lesions on diffusion-weighted imaging in TIA patients. Stroke. 2009;40:2229–2232. doi: STROKEAHA.108.537969 [pii] 10.1161/STROKEAHA.108.537969

24. Lodha N, Patel P, Harrell J, Casamento-Moran A, Zablocki V, Christou EA, Poisson SN. Motor impairments in transient ischemic attack increase the odds of a positive diffusion-weighted imaging: A meta-analysis. Restor Neurol Neurosci. 2019;37:509–521. doi: 10.3233/RNN-190940

